# Antibody Immunological Imprinting on COVID-19 Patients

**DOI:** 10.1101/2020.10.14.20212662

**Authors:** T Aydillo, Alexander Rombauts, Daniel Stadlbauer, Sadaf Aslam, Gabriela Abelenda-Alonso, Alba Escalera, Fatima Amanat, Kaijun Jiang, Florian Krammer, Jordi Carratala, Adolfo García-Sastre

## Abstract

While the current pandemic remains a thread to human health, the polyclonal nature of the antibody response against SARS-CoV-2 is not fully understood. Other than SARS-CoV-2, humans are susceptible to six different coronaviruses, and previous exposure to antigenically related and divergent seasonal coronaviruses is frequent. We longitudinally profiled the early humoral immune response against SARS-CoV-2 on hospitalized COVID-19 patients, and quantify levels of pre-existing immunity to OC43, HKU1 and 223E seasonal coronaviruses. A strong back-boosting effect to conserved, but not variable regions of OC43 and HKU1 betacoronaviruses spike protein was observed. All patients developed antibodies against SARS-CoV-2 spike and nucleoprotein, with peak induction at day 7 post hospitalization. However a negative correlation was found between antibody memory boost to human coronaviruses and induction of IgG and IgM against SARS-CoV-2 spike. Our findings provide evidence of immunological imprinting that determine the antibody profile to COVID-19 patients in an original antigenic sin fashion.

---

Since January 20202, the Severe Acute Respiratory Syndrome Coronavirus 2 (SARS-CoV-2) virus has been spreading globally causing the first documented pandemic of coronavirus in history (*1, 2*). SARS-CoV-2 is a betacoronavirus that belongs to a large family of viruses capable to infect both mammals and birds. Humans are susceptible to six other six viruses from the genus alpha- and beta-coronavirus (*3*). All of them typically cause respiratory illness but to a different extent. While Severe Acute Respiratory Syndrome Coronavirus 1 (SARS-CoV-1) and Middle East Respiratory Syndrome Coronavirus (MERS-CoV), are highly pathogenic betacoronaviruses that caused zoonotic outbreaks in humans in the last 20 years (*4, 5*), the alphacoronaviruses 223E and NL63, and the betacoronaviruses OC43 and HKU1, frequently cause mild upper respiratory disease and have been circulating in humans for at least 100 years (*3, 6*). The ongoing pandemic of COVID-19, the disease caused by SARS-CoV-2, is still challenging healthcare systems and the research community. SARS-CoV-2 can cause a different range of clinical manifestations, from asymptomatic to severe respiratory syndrome. However a high percentage of severe cases have also been reported and estimated numbers of patients that succumbed to COVID-19 disease are more than a million according to WHO as October 2020 (*2, 7*) (https://covid19.who.int/). While no vaccine is available yet, at least three candidates are already on phase 3 clinical trials in the US (*8-10*). Previous studies have shown that a main target of antibody responses to coronaviruses is the spike, the surface glycoprotein that mediates attachment to the host receptor, ACE2, and membrane fusion (*11, 12*). Moreover, antibodies directed against the receptor binding domain (RBD) of the spike, are known to neutralize the virus (*13, 14*). RBD antibodies are highly specific and in general do not cross-react among the seasonal human coronavirus (*15-18*). In addition, the more cross-reactive viral nucleoprotein (NP) has also shown to be immunogenic and induce antibodies in COVID-19 patients (*11, 18, 19*). However, in contrast to RBD antibodies, NP antibodies are not able to neutralize the virus in tissue culture.

While some cell-mediated and serum cross-reactivity between epitopes from SARS-CoV-2 and seasonal human coronaviruses has been demonstrated (*18, 20, 21*), whether previous exposure to other human coronaviruses can influence the immunological outcome while encounter a novel but closely related antigen is not clear. This phenomenon termed immune imprinting or original antigenic sin, refers to the preference of the immune system to recall existing memory responses, rather than stimulating de novo responses (*22*). This has been well studied for viruses like influenza (*23, 24*), and is a fundamental piece to inform vaccine development (*25, 26*). In here, we profiled the antibody responses of a longitudinal cohort of hospitalized patients with COVID-19 disease. We characterized antibody responses against both SARS-CoV-2 and selected seasonal coronavirus proteins being targeted by the humoral immune system and explore the role of immunological imprinting on COVID-19 patients’ immune response.

## RESULTS

### THE BACO COHORT

Thirty-seven COVID-19 patients were recruited at the University Hospital of Bellvitge during the first wave of SARS-CoV2 in Barcelona (Spain) from March 26^th^ to May 28^th^, 2020. Mean age was 65 years and 67% were male. Chronic comorbidities were frequent among COVID-19 patients (25, 67.7%). In particular, In particular, 16 (43.2%) of patients were obese (body mass index, BMI>30) at time of hospitalization. A high percentage of patients had respiratory symptoms such as coughing (26, 70.3%) and dyspnea (14, 37.8%), whereas diarrhea was also present in 7 (18.9%) of thevpatients. While no remdesivir was available, lopinavir/ritonavir was used for 17 (45.9%) patients. All patients, except one (36, 97.3%), developed SARS-CoV-2 viral pneumonia and 4 (10.8%) required intensive care unit (ICU) admission. Five (13.5%) patients died. Demographics, clinical characteristics, interventions such as drug therapy and outcomes are detailed in Table 1.

**Table 1.** Demographics and clinical characteristics of the BACO Cohort.

|  | **Total (n=37)** |
| --- | --- |
| **Demographics and Comorbidities** |  |
| Age (mean, IQR) | 67 (25) |
| Men (n, %) | 25 (67.6) |
| Co-morbidities (n, %) | 25 (67.7) |
| Lung Disease (n, %) | 7 (18.9) |
| Diabetes mellitus | 7 (18.9) |
| Heart disease (n, %) | 5 (13.5) |
| Kidney disease (n, %) | 3 (8.1) |
| Obesity (n, %) | 16 (43.2) |
| SOTR (n, %) | 1 (2.7) |
| **Signs and Symptoms** |  |
| Days from symptom onset to enrollment (mean, range) | 7.19 (2-14) |
| Days of fever (mean, range) | 4.68 (0-12) |
| Throat ache (n, %) | 4 (10.8) |
| Cough (n, %) | 26 (70.3) |
| Dyspnea (n, %) | 14 (37.8) |
| Diarrhea (n, %) | 7 (18.9) |
| SpO2<94% (n, %) | 14 (37.8) |
| **Drug Therapy** |  |
| Hydroxychloroquine (n, %) | 36 (97.3) |
| Lopinavir/Ritonavir (n, %) | 17 (45.9) |
| Tocilizumab (n, %) | 10 (27) |
| Antibiotics (n, %) | 19 (51.4) |
| Corticosteroids (n, %) | 18 (48.6) |
| **Outcomes** |  |
| Pneumonia (n, %) | 36 (97.3) |
| ICU (n, %) | 4 (10.8) |
| Days from hospitalization to ICU (mean, range) | 9.5 (5-12) |
| Days in ICU (mean, range) | 15 (15-22) |
| Non-mechanical Ventilation (n, %) | 11 (29.7) |
| Mechanical Ventilation (n, %) | 2 (5.4) |
| Nosocomial Co-Infection (n, %) | 2 (5.4) |
| Mortality (n, %) | 5 (13.5) |
| Days of hospitalization (mean, range) | 11.2 (2-47) |

SOTR: Solid Organ Transplant Recipient; SpO2<94%: pulse oximetry below 94%; ICU: intensive care unit

Acute blood samples were collected longitudinally in the BACO cohort at the recruitment upon hospital admission, and at day 3 and 7 in 33 (89.1%) and 22 (59.4%) patients, respectively. Whereas more than half of the patients (25, 67.5%) were recruited within the first week of symptom onset, 12 (32.4%) patients had longer periods until hospitalization. COVID-19 survivors were followed up in the convalescence period and 28 out of 32 survivors (87.5%) had a another blood draw after hospital discharge with a mean time of 46 days post-recruitment (range, 30-56 days).

### COVID-19 PATIENTS DEVELOPED ANTI SARS-CoV-2 ANTIBODIES LINKED TO BACK-BOOSTING OF ANTIBODIES AGAINST S2 DOMAIN OF BETA CORONAVIRUSES

To profile the early antibody response on COVID-19 patients we investigated levels of neutralizing antibodies against authentic SARS-CoV-2 virus and IgG/IgM enzyme-linked immunosorbent assays (ELISAs) against multiple antigens including the full length spike (S), the spike receptor binding domain (RBD S) and the nucleoprotein (NP) of SARS-CoV-2 (Fig. 1A). All patients developed detectable levels of neutralizing antibodies at day 7 post-recruitment while levels remained stable during the convalescence phase, except for two survivors. Similar responses were found by ELISA assays, although higher levels of antibodies against IgG S compared to IgG RBD were present. While comparing to the induction of anti-IgM, IgG isotype reached higher titers than IgM anti-spike; whereas anti-NP protein IgG overlapped with the induction of the anti-S IgG. Although the S gene of SARS-CoV-2 is highly divergent to other human coronaviruses (from now hCoV) (*6*), infection of hCoV among humans is frequent (*3, 27*), causing mild respiratory disease. Given the high probability of previous exposure to seasonal coronaviruses, we screened levels of antibodies against selected human alpha- (223E) and beta-coronaviruses (HKU1, OC43). Antigens tested included full length S protein for all three hCoVs together with the HKU1 RBD S (Fig. 1B). COVID-19 patients exhibited an outstanding back-boosting of antibodies to the beta-hCoV spikes, with similar increase that the one observed for SARS-CoV-2 spike and neutralizing titers. Although IgG levels against 223E were already high at baseline, no increase was detected at any time point during the follow up on patients with COVID-19. Interesting, no back-boosting was found when we tested antibody titers against the more divergent S1 domain of HKU1, pointing to an increase of immune responses towards conserved epitopes of the S2 domain of the spike protein of beta-hCoV. Similar as influenza viruses, HKU1 and OC43 use sialic acids as canonical receptor to infect human cells (*28*). This is mediated by an additional surface protein in these viruses with hemagglutination activity (HE protein) No increase in OC43 hemagglutination inhibitory antibodies was found in COVID-19 patients, consistent with the lack of HE in SARS-CoV-2. We next tested the correlation between neutralization activity and levels of anti-SARS-CoV-2 antibodies. Scatterplot matrices shown in Fig. 2A indicates that the antibodies detected against SARS-CoV-2 antigens correlated well with neutralizing activity, with Pearson R^2^ ranging from 69% to 81%, in the particular case of IgG against the RBD S of SARS-CoV-2.

**Fig. 1.**
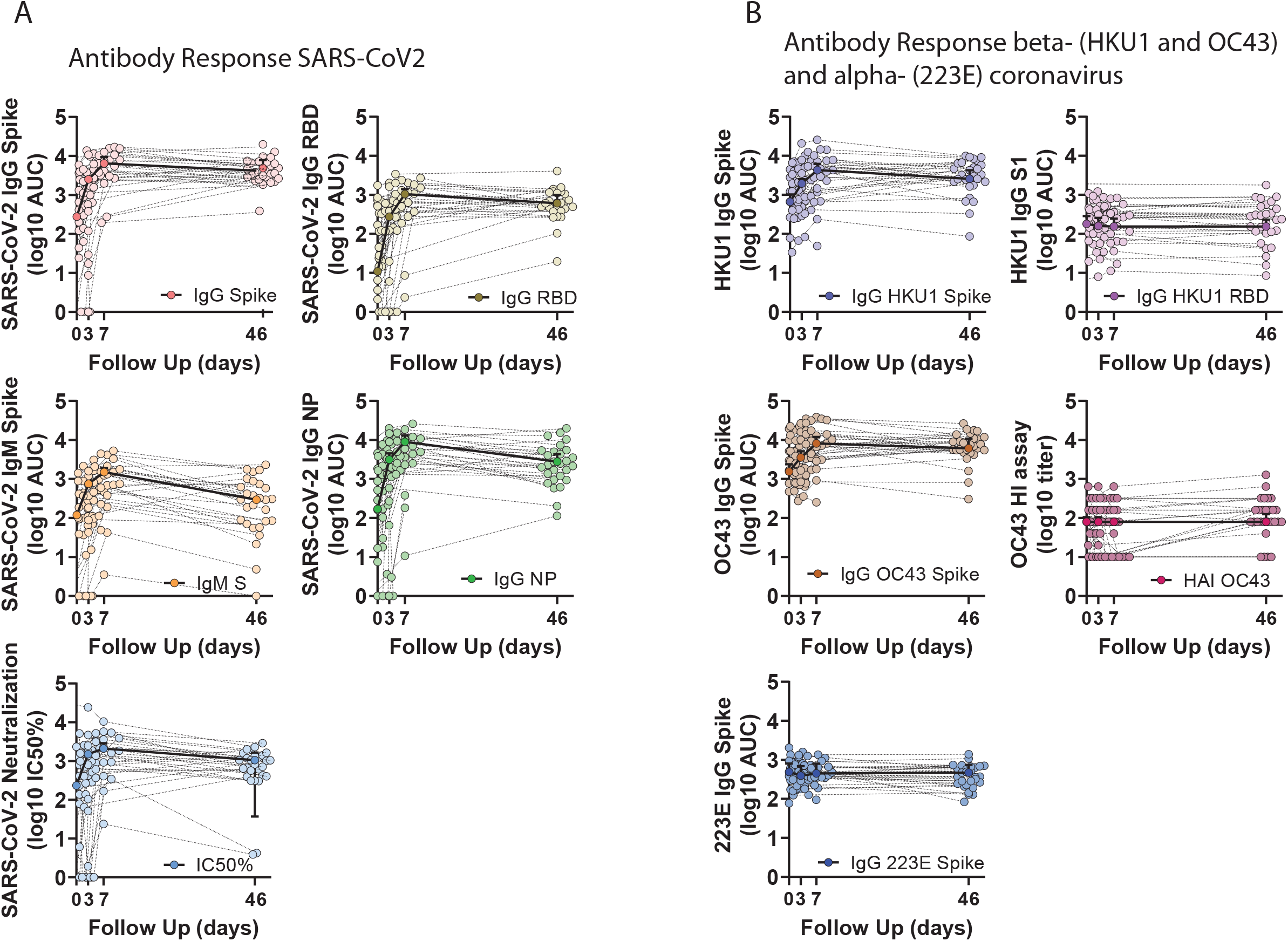
Longitudinal profile of antibody responses to SARS-CoV-2 antigens and selected seasonal human coronaviruses. Serum from hospitalized COVID-19 patients was analyzed at baseline, at hospital recruitment and day 3 and 7. A subsequent sample was collected in the convalescence period in the COVID-19 survivors with mean time of 46 days. A) ELISA AUC titers against different antigens of SARS-CoV-2 at each time point: IgG spike, IgG RBD, IgM spike and IgG NP. Neutraliz-ing titers were also performed and IC50% at each time point is also shown. B) ELISA AUC titers against antigens from seasonal coronaviruses: IgG HKU1 spike, IgG HKU1 S1, IgG OC43 spike and IgG 223E. Hemagglutination inhibition assay were also performed for OC43 and end point titers are shown at each time point. Median and CI 95% is shown

**Fig. 2.**
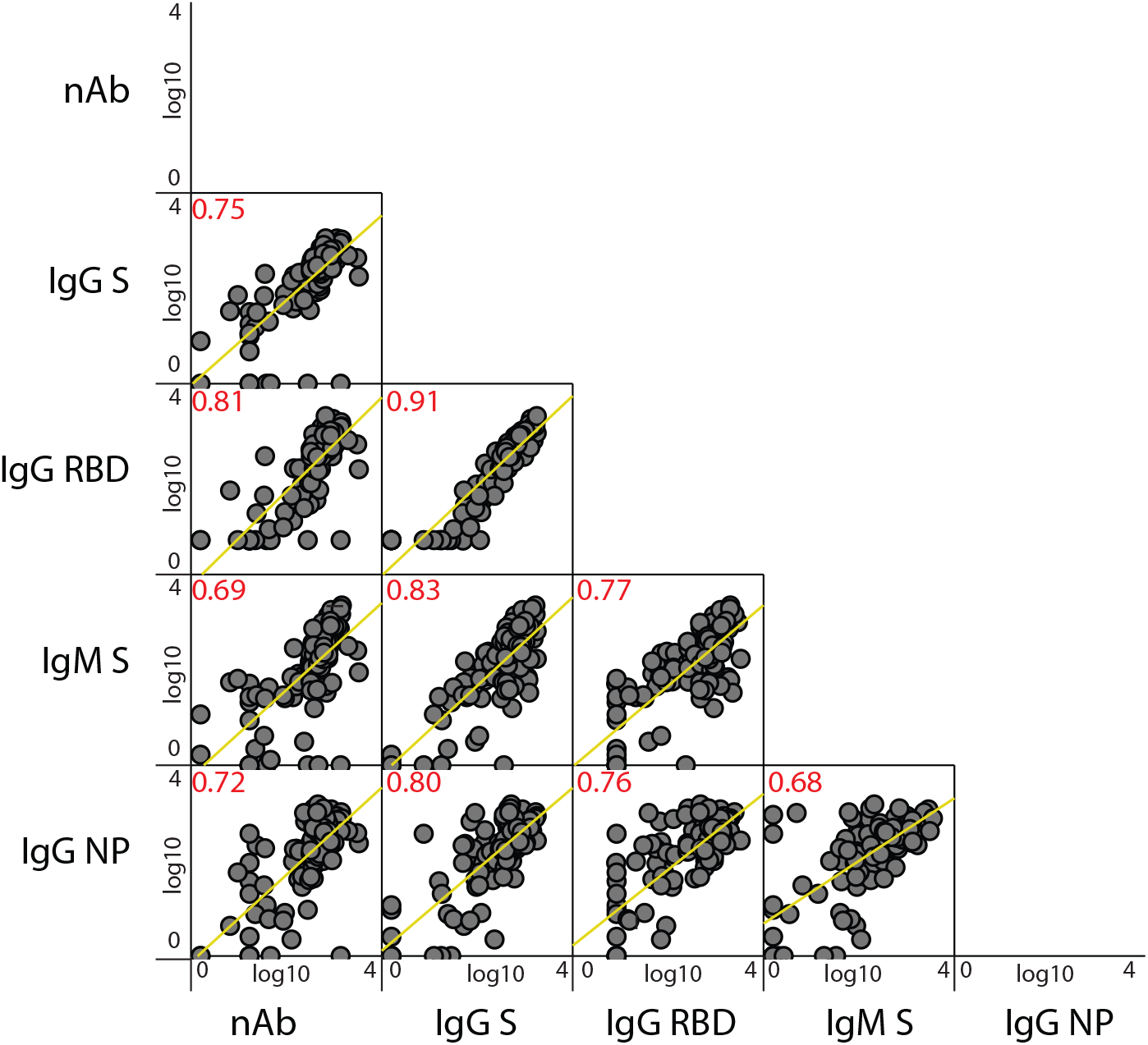
Scatterplot matrix of the relationship between measured SARS-CoV-2 antibody responses in the COVID-19 patients. Relationship of each antibody levels is tested for the correlations shown: ELISA AUC titers against SARS-CoV-2 (IgG spike, IgG RBD, IgM spike and IgG NP) and neutralizing titer (nAb, IC50%). Pearson coefficient of statistically significant correlations are indicated in red. Fitted linear regression lines for each interaction are shown in yellow. Matrix axis are log10 values scaled from 0 to 4.

### IMMUNOLOGICAL IMPRINTING RESULTS IN A BIAS IN THE INDUCTION OF ANTIBODIES TO CONSERVED VERSUS VARIABLE REGIONS OF THE SARS-COV-2 SPIKE

Given the strong back-boosting observed for the conserved epitopes of the S domains of human betacoronaviruses in patients with COVID-19, we next investigated whether a strong back-boosting might reduce the induction of *de novo* humoral immune responses against specific epitopes of the spike of SARS-CoV-2. To test this hypothesis we defined *de novo* antibody responses and determined the average fold-induction at day 3, 7 and convalescence (in the survivors group) over baseline levels at the recruitment in the study. Overall, all patients had high induction of SARS-CoV-2 S and RBD antibodies at day 7 post recruitment. IgG titers against the S and RBD of SARS-CoV-2 remained stable at the convalescence time point with similar levels compared to day 7. By contrast, IgM against the S, IgG against NP and neutralizing titers against the authentic SARS-CoV-2 virus decreased to levels resembling those at day 3 (Fig. 3A). Median fold-induction and pairwise comparisons including adjusted p values between each time points and their association over time are shown in Fig.3A. Comparable results were found for induction of antibodies against the spike of seasonal betacoronaviruses (Supplemental Fig. 1).

**Fig 3.**
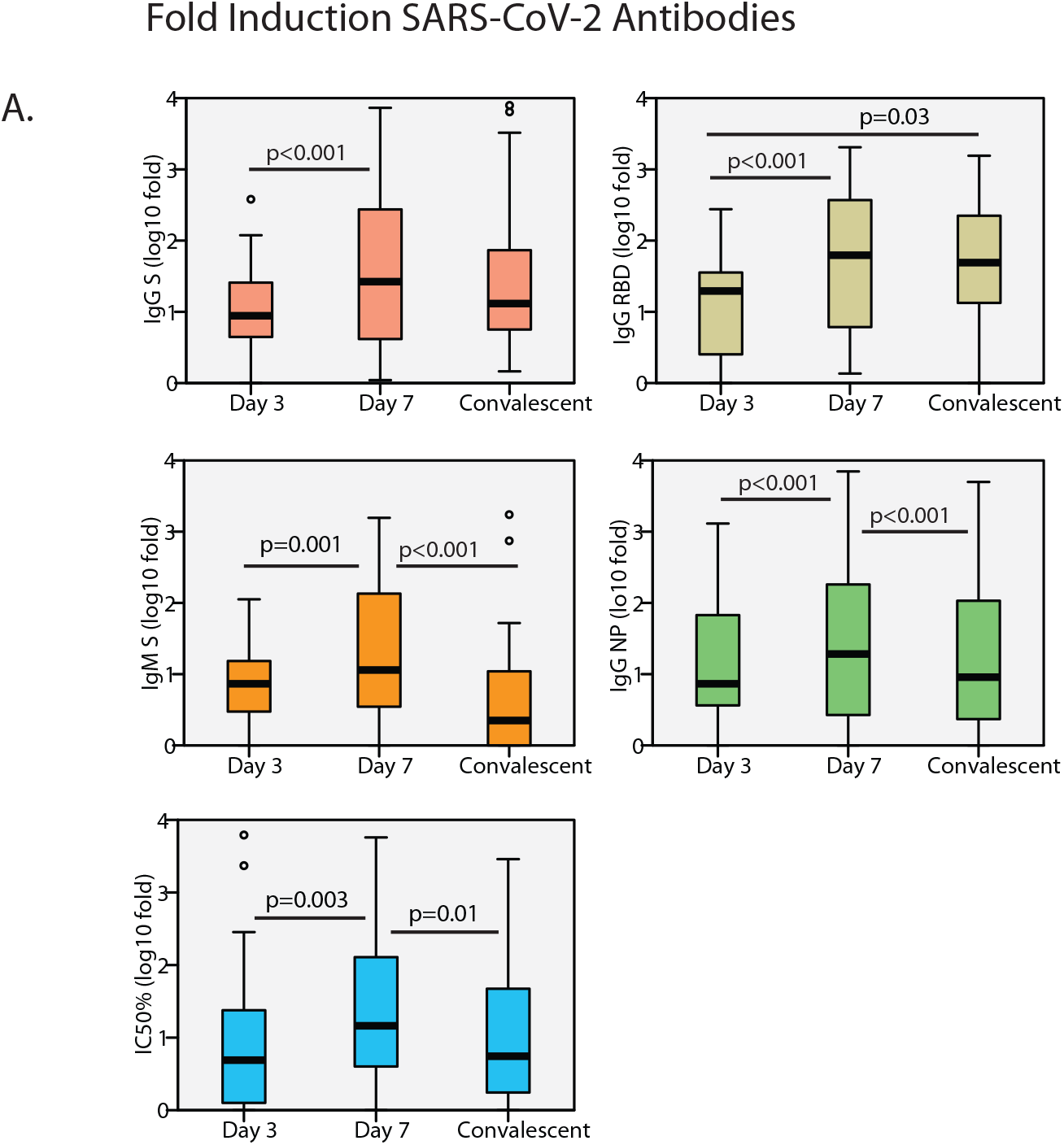
Fold induction of anti-SARS-CoV-2 antibodies over time. A) Boxplot diagram of fold induction values of ELISA AUC titers against SARS-CoV-2 at each time point: IgG spike, IgG RBD, IgM spike and IgG NP; and neutralizing titer (IC50%). Related-samples Friedman’s two-way comparison was performed and significant adjusted p values for pairwise comparisons are shown for each antibody levels at each time point. Black bar indicated median values, box indicates IQR (Q1-Q3), lines indicate minimun and maximun. Outliers from the observed distribution are shown when present in each case.

We next examined the relationship between pre-exposure to HKU1 and OC43 viruses and induction of SARS-CoV-2 S and RBD antibodies in our cohort. Pearson correlation matrices were used to test whether pre-existing immunity against cross-reactive betacoronaviruses can influence subsequent immune response to SARS-CoV2 S and NP antigens. We leveraged on the IgG levels at baseline against seasonal human coronaviruses and split the analysis by virus subtype (Fig. 4). Striking differences were found according to viruses lineages. While pre-existing IgG levels against HKU1 and OC43 spike protein negatively impacted the induction of *de novo* IgG and IgM against SARS-CoV-2 antigens (Fig. 4A-B), no influence was found when tested the relationship between pre-existing anti-223E IgG levels (Fig. 4D). Moreover, correlations became stronger over time, and while this correlation was lower at day 3, a stronger correlation was found at day 7, and convalescence time points in the survivor patients. Besides, comparable performance was observed when testing the subsequent induction of the IgG antibodies against the variable RBD domain of SARS-CoV-2 spike suggesting that pre-existing immunity against seasonal beta coronavirus bias the humoral response towards beta-coronaviruses cross-reactive antibodies in detriment of antibodies against the more divergent antigenically unique domains of the S of SARS-CoV-2, such as those of the RBD domain (Fig. 4A-B). This was also evidence by the lack of impact of pre-existing IgG levels of HKU1 S1, which RBD is antigenically different from to the RBD of SARS-CoV-2, on specific SARS-CoV-2 antibodies induction (Fig. 4C). Thus, only the levels of antibodies against cross-reactive epitopes of human betacoronaviruses had an effect on the subsequent antibody response to SARS-CoV2 unique spike antigens. Because neutralization activity has been linked to in vivo protection after challenge with SARS-CoV-2 (*29*), we also tested if immune imprinting could hinder the induction of neutralizing antibodies against SARS-CoV-2. Although not statistically significant, linear regression analysis determined a standardized beta coefficient of −0.32 (95% CI −0.35 to 0.05, p=0.13) and −0.31 (95% CI −0.28 to 0.02, p=0.1) at day 7 and convalescence time points, respectively, for HKU1 spike pre-existing levels approximating a negative impact of HKU1 pre-existing immunity in induction of neutralizing antibodies against SARS-CoV-2 in COVID-19 patients (Fig. 5A). A similar trend was found for the levels of pre-existing antibodies against OC43 spike (Fig. 5B). Scatterplots and the predicted regression lines for the relationship of induction of antibodies with neutralizing activity and pre-exposure to betacoronaviruses are shown in Fig. 5A-D according to time points.

**Fig 4.**
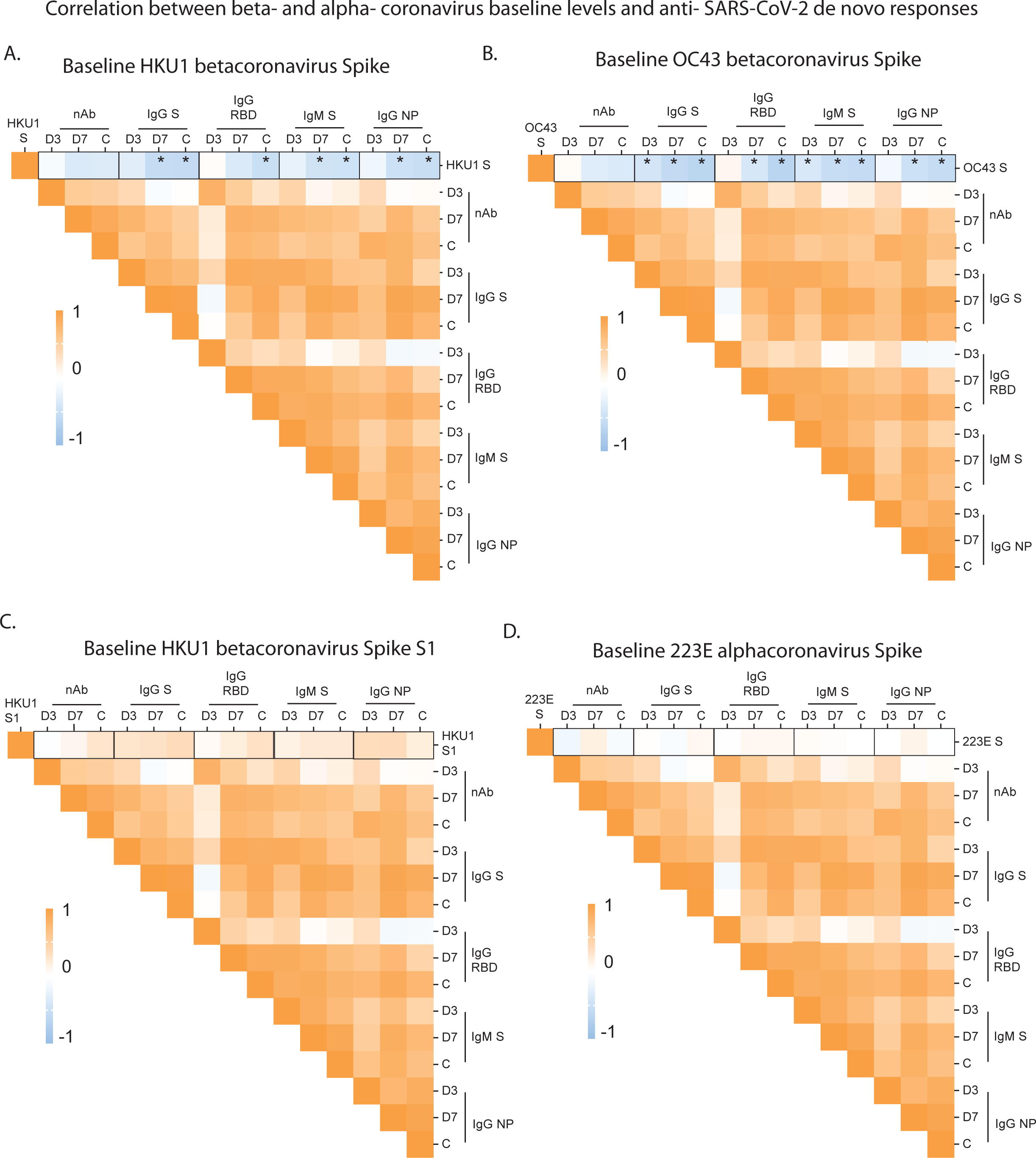
Immunological imprinting on SARS-CoV-2 humoral response. A-B) Heatmap of Pearson correlation matrices between pre-existing levels of seasonal hCoV: IgG HKU1 S, IgG HKU1 S1, IgG OC43 S and IgG 223E; and fold induction of SARS-CoV-2 antibodies at each time point: neutralizing (nAb), IgG spike, IgG RBD, IgM spike and IgG NP. Statistically significant correlations in the underlined intersections are indicated with asterisk (*); D3: day 3; D7: day 7; C: convalescence.

**Fig. 5.**
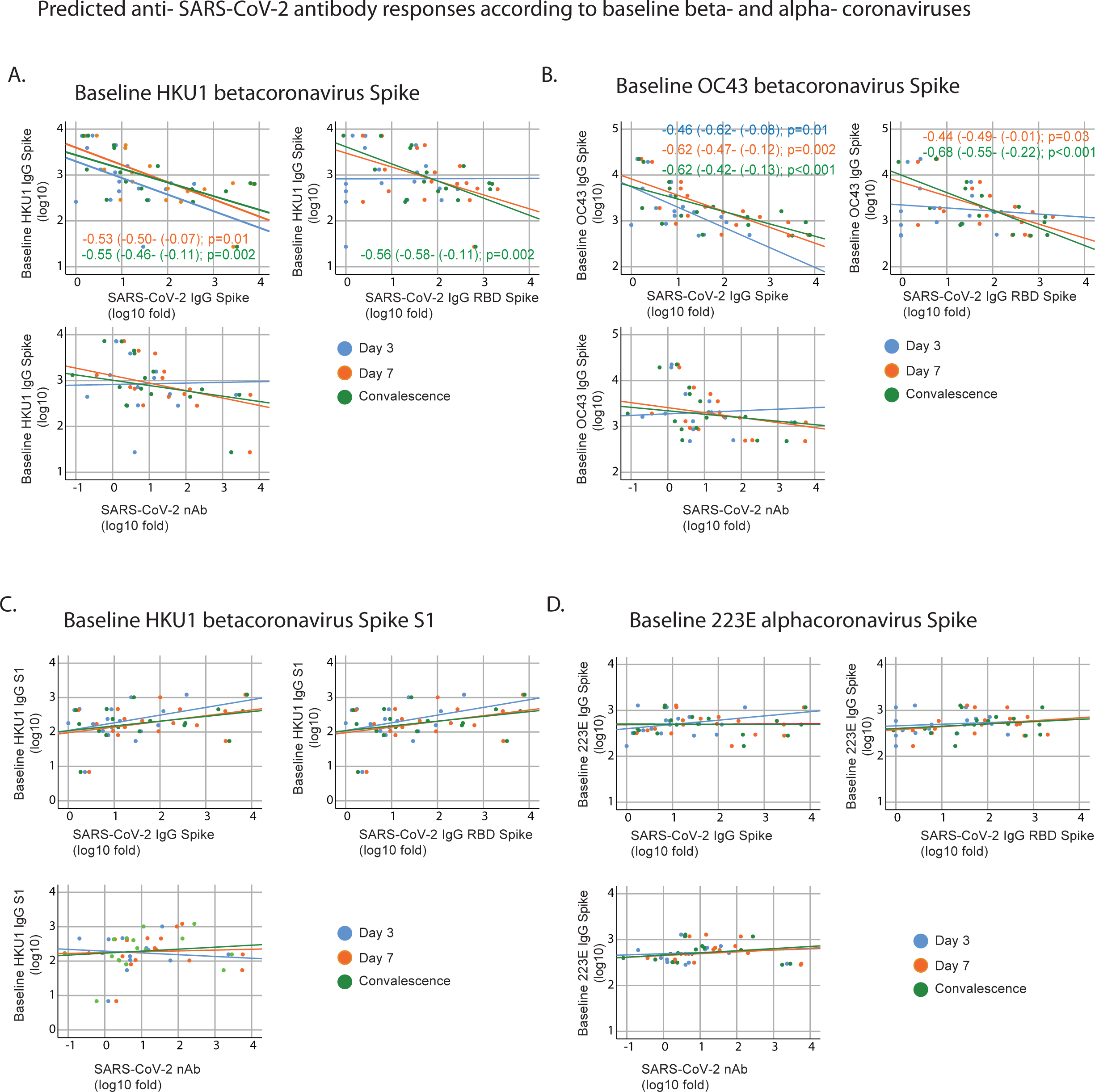
Cross-reactivity with conserved epitopes against selected betacoronavirus predicts negative influence on de novo anti SARS-CoV-2 antibody responses. A-D) Scatterplot of baseline IgG levels for HKU1, OC43 and 223E S protein; and HKU1 S1 and fold induction of SARS-CoV-2 antibodies: neutralizing (nAb), IgG spike, IgG RBD. Overlay shows relationship with induction of de novo antibodies against SARS-CoV-2 at each time point. Fitted linear regression and standardized beta coefficient (95% Confidence Interval, CI) for significant linear regressions are shown.

Since levels of cross-reactive antibodies and back-boosting may differ across patients, we normalized levels of IgG against seasonal human coronavirus antigens by the levels of anti-spike IgG from SARS-CoV-2 virus at the same time points; and tested whether those patients with higher hCoV/SARS-CoV-2 IgG ratio had lower induction of neutralizing antibodies. After linear regression analysis some disparities were found (Supplemental Fig. 2). In general, the higher hCoV/SARS-CoV-2 IgG ratio for HKU1 and OC43 IgG S at baseline and day 3, the lower was the induction of antibodies with neutralizing activity to SARS-CoV-2, suggesting some limitations for the ability to elicit robust protective antibody responses against novel antigenic epitopes of SARS-CoV-2 in patients with high levels of cross-reactive antibodies against circulating beta-coronaviruses.

## DISCUSSION

Our findings provide a dynamic characterization of the antibody response against SARS-CoV-2 in COVID-19 patients and provide evidence of immune imprinting in these patients. Our results demonstrate back-boosting in the BACO cohort against the conserved epitopes of the spike protein of OC43 and HKU1 betacoronaviruses, as no induction was detected for the variable regions of these viruses, such as the S1 domain, or to more divergent seasonal alpha-coronaviruses such as 223E. Although antibody cross-reactivity has been reported in cross-sectional studies (*17, 18, 20, 21*), our cohort has allowed for quantification and detailed representation of the longitudinal outcome of the immune response by taking into consideration past exposure to related antigens.

Neutralization activity of antibodies might be used as a proxy for protection against SARS-CoV-2 infection (*30, 31*). IgG RBD and spike SARS-CoV-2 responses showed persistence over the time period of our study with slight changes in antibody levels in convalescent sera as compared to the peak of antibody induction at day 7. Importantly, other betacoronavirus spikes, like HKU1 and OC43, while not bearing neutralizing of authentic SARS-CoV-2 activity in vitro, limited the induction of *de novo* responses for all the SARS-CoV-2 antigens tested. All patients also developed detectable levels of spike IgG/IgM and NP IgG. Although not significant correlation was found between pre-exposure to seasonal coronaviruses and induction of protective antibodies with neutralization activity, simple linear regression estimated a negative relationship, and the predicted line approximated a negative influence on development of de novo neutralizing antibodies over time. Equivalent, baseline levels of HKU1 or OC43 spike after SARS-CoV-2 IgG levels normalization conditioned day 3 induction of neutralizing antibody levels.

Our observation has important implications on the development of COVID-19 vaccines and the potential interactions with pre-existing immunity should be taken into consideration in the path to an effective vaccine, to ultimately control the ongoing pandemic. Most of the COVID-19 vaccines are based on full length S protein of SARS-CoV-2 (*32*), which is known to contain cross-reactive non-neutralizing epitopes common with seasonal human betacoronaviruses. Importantly, we still do not know if such non-neutralizing antibodies in vitro contribute to protection or disease in vivo by beneficial activities such as antibody-mediated cytotoxicity, or by detrimental activities, such as antibody dependent enhancement disease. In any case, our results demonstrate that the antibody response against SARS-CoV-2 infection and, potentially vaccination, is influenced by imprinting of the B cell compartment due to previous exposure to seasonal human betacoronaviruses. It will be important to investigate the potential functional consequences of this imprinting in the induction of protective immune responses after SARS-CoV-2 infection and vaccination on the long term.

## Data Availability

All data is available in the manuscript or the supplementary materials. Serum samples are available upon reasonable request from Dr. Garcia-Sastre under a material agreement with Icahn School of Medicine at Mount Sinai. Reagents used are almost exclusively commercially available and non-proprietary.

## ACKNOWLEDGMENTS

We thank the BACO cohort patients and their families. We also thank Randy Albrecht for support with the BSL3 facility and procedures at the ISMMS and Richard Cadagan for excellent technical assistance. We also want to thank Kizzimekia Corbett and Barney Graham at the Vaccine Research Center at NIAID for sharing expression plasmids for hCoV spike proteins.

## Funding

This work was partly funded by CRIP (Center for Research on Influenza Pathogenesis), a NIAID funded Center of Excellence for Influenza Reserch and Surveillance (CEIRS, contract #HHSN272201400008C), by SEM-CIVIC, a NIAID funded Collaborative Influenza Vaccine Innovation Center (contract #75N93019C00051), and by the generous support of the JPB Foundation, the Open Philanthropy Project (research grant 2020-215611 (5384)) and anonymous donors to AG-S.

## Author contributions

TA performed experiments, analyzed data and wrote the manuscript. AR, GAA and JC collected samples and data. DS, SA, AE, KJ and FA performed experiments. FK provided reagents, methods and expertise. TA and AGS conceived and designed the study. TA, JC and AGS supervised the study. All of the authors reviewed and edited the manuscript.

## Competing Interests

AG-S is inventor of patents owned by the Icahn School of Medicine at Mount Sinai in the field of influenza virus vaccines. The AG-S lab has received research funds from Avimex, GSK and 7Hills to investigate novel influenza virus vaccines. FK is a named as inventor on IP covering serological assays and vaccines for SARS-CoV-2 which have been filed by the Icahn School of Medicine at Mount Sinai. Other authors declare no competing interests.

## Data and materials availability

All data is available in the manuscript or the supplementary materials. Serum samples are available upon reasonable request from Dr. García-Sastre under a material agreement with Icahn School of Medicine at Mount Sinai. Reagents used are almost exclusively commercially available and non-proprietary.

## EXPERIMENTAL MODEL AND SUBJECT DETAILS

### THE BACO COHORT

An observational prospective human cohort study of subjects with COVID-19 disease was carried out during the first pandemic wave (March-May 2020) of SARS-CoV-2 in Barcelona (Spain): BACO Cohort. A positive case was defined according to international guidelines when nasopharyngeal (NP) swab tested positive for SARS-CoV2 by real-time polymerase chain reaction (RT-qPCR) upon hospital admission. All patients or their legally authorized representatives provided informed consent. Serum and samples were collected at the enrollment in the study (baseline), and at day 3 and 7 post-enrollment. A convalescence sample was collected on survivors after recovery and hospital discharge with a mean time of 46 days (range, 30-56 days). Data on demographics, including age and sex, co-morbidities, clinical signs and symptoms, interventions, and outcomes are described in Table 1. The study protocol was approved by the Institutional Review Board of University Hospital of Bellvitge, Barcelona, Spain; and by the Icahn School of Medicine at Mount Sinai, New York, US.

### CELL LINES

Vero E6 cells were originally purchased from the American Type Culture Collection (ATCC, Cat# CRL-1586). Cells were maintained in Dulbecco’s modified Eagle’s medium (DMEM) w/ L-glutamate, sodium pyruvate (Corning) supplemented with 10% fetal bovine serum (FBS), 100 U penicillin per ml, and 100 mg streptomycin per ml. HCT-8 human cells line was obtained from the ATCC (Cat#CCL-24) and maintained in Roswell Park Memorial Institute (RPMI) 1640 medium (Gibco) supplemented with 10% fetal bovine serum (FBS), 100 U penicillin per ml, and 100 mg streptomycin per ml. Cell lines were supplemented with Normocyn (Invivogen, Cat. ant-nr-1) to prevent Mycoplasma contamination.

### VIRUS STRAINS

SARS-CoV-2, Isolate USA-WA1/2020 was initially obtained from BEI Resources (Cat#NR-52281) and further propagated in Vero E6 cells as previously described (20). Human coronavirus OC43 was obtained from the ATCC (Cat#VR-1558) and propagated on HCT-8 cells following ATCC recommendations.

## METHODS DETAILS

### MICRONEUTRALIZATION ASSAYS

Micro neutralization (MN) assay for antibody characterization was performed as described previously (20) with modifications. Briefly, Vero E6 cells were seeded in a 96-well cell culture plate with complete Dulbecco’s Modified Eagle Medium (cDMEM)(Corning) [(Penicillin-streptomycin (Corning), non-essential amino acids (Corning), 10% fetal bovine serum (FBS) (Peak)]. The following day, heat-inactivated serum samples were serially diluted three-fold in 1xminimum essential medium (MEM) with 2% FBS with a final volume of 200μl. 80μl of serum dilution was transferred to a new 96-well plate and 600 TCID50/well of SARS-CoV-2 (80μl/well) and mixed with serum dilution and incubated for 1hr at 37°C. Then, cDMEM was removed from Vero e6 cells and 120μl of virus-serum mixture was added to the cells. The cells were incubated at 37°C for 1 hour. Virus-serum mixture was removed from the cells and 100μl of serum dilutions and 100μl of 1xMEM with 2% FBS was added to the cells. The cells were incubated for 24hours and then fixed with 10% paraformaldehyde (Polysciences) for 24 hours at 4°C. Following fixation, the cells were washed with phosphate-buffered saline (Corning) with tween-20 (Fisher) (PBST) and permeabilized with 0.1% Triton X-100 (Fisher) for 15 min at room temperature. The cells were washed three times using PBST and blocked with 3% milk in PBST for 1 hour at room temperature. Then, the cells were incubated with mAB 1C7 (anti-SARS nucleoprotein antibody, kindly provided by Dr. Moran) at a dilution of 1:1000 in 1% milk in PBST and incubated for 1hr at room temperature. The cells were washed three times with PBST. Then, the cells were incubated with goat anti-mouse IgG-HRP (Abcam) at a dilution of 1:10000 in 1% milk in PBST and incubated for 1hr at room temperature. The cells were washed three times with PBST and TMBE Elisa peroxidase substrate (Rockland) was added. After 15 min incubation, sulfuric acid 4.0N (fisher) was added to stop the reaction and the readout was done using a Synergy H1 plate reader (BioTek) at an OD450.

### RECOMBINANT PROTEINS

The recombinant spike protein and recombinant RBD of SARS-CoV-2 were generated and expressed as previously described in detail (21). In brief, the mammalian cell codon-optimized nucleotide sequence for the soluble version of the spike protein (amino acids 1-1,213) including a C-terminal thrombin cleavage site, signal peptide, hexahistidine tag and T4 foldon trimerization domain were cloned into pCAGGS mammalian expression vector. The sequence of the spike protein was additionally modified to remove the polybasic cleavage site and two proline residues introduced to increase protein stability. The nucleotide sequence for the RBD (amino acids 319-541) including a signal peptide was cloned into pCAGGS. The expression plasmids encoding for the spike of common human coronavirus 229E, OC43 and HKU1, and the expression plasmid encoding for SARS-CoV-2 NP were obtained from the NIH (Kizzmekia Corbett and Barney Graham). The recombinant proteins were expressed in Expi293F cells (Thermo Fisher) using the ExpiFectamine 293 Transfection Kit (Thermo Fisher) according to manufacturer’s protocols. Cell supernatant was harvested and the proteins purified using Ni-NTA Agarose (Qiagen). The proteins were concentrated in Amicon centrifugal units (EMD Milipore) and correct size confirmed by reducing sodium dodecyl sulfate-polyacrylamide gel electrophoreses (SDS-PAGE). The recombinant S1 subunit of HKU1 was purchased from Sino Biological (Cat. 40021-V08H).

### ENZYME-LINKED IMMUNOSORBENT ASSAY (ELISA)

Ninety-six-well microtiter plates (Thermo Fisher) were coated with 50 μL recombinant protein (RBD, SARS-CoV-2 full-length spike, SARS-CoV-2 NP, OC43 spike, 229E spike or HKU1 spike respectively) at a concentration of 2 ug/mL overnight, 4°C. The next day, the plates were washed three times with PBS (phosphate-buffered saline; Gibco) containing 0.1% Tween-20 (T-PBS, Fisher Scientific) using an automatic plate washer (BioTek). After washing, the plates were blocked for 1h at room temperature with 200 ul blocking solution (PBS-T with 3% (w/v) milk powder (American Bio)) per well. The blocking solution was removed and serum samples diluted to a starting concentration of 1:80, serially diluted 1:3 in PBS-T supplemented with 1% (w/v) milk powder and incubated at room temperature for 2 h. The plates were washed three times with PBS-T and 50 ul anti-human IgG (Fab-specific) horseradish peroxidase antibody (HRP, Sigma, #A0293) diluted 1:3,000 in PBS-T containing 1% milk powder was added to all wells and incubated for 1 h at room temperature. The plates were washed three times using the plate washer and 100 μL SigmaFast o-phenylenediamine dihydrochloride (OPD; Sigma) was added to all wells for 10 minutes. The enzymatic reaction was stopped with 50 μL 3M hydrochloric acid (Thermo Fisher) per well and the plates read at a wavelength of 490 nm with a plate reader (BioTek). The results were recorded in Microsoft Excel and the endpoint titer and area under the curve values calculated in GraphPad Prism.

### HEMAGGLUTINATION INHIBITION (HAI) ASSAY

Serum samples were incubated overnight with receptor-destroying enzyme (RDE; Denka Seiken) for 16-18 h in a 37°C water bath. Three volumes (relative to serum) of 2.5% sodium citrate solution were added and RDE were heat inactivated at 56°C in a water bath (30 minutes). Final serum dilutions were adjusted to 1:10 in PBS. OC43 virus was diluted to a final concentration of 8 HA units/50 μL in Fluorescent Treponemal Antibody (FTA) hemagglutination (HA) buffer (BD Biosciences). Two-fold dilutions of RDE treated serum (25 μL) were incubated with equal amount of the virus at 8 HA units/50 μL (30 minutes, room temperature). Chicken red blood cells (RBCs) (Lampire Biological) at 0.5% in HA buffer (50 μL) were added and incubated 45 minutes at 4°C. The HAI titer was determined by taking the reciprocal dilution of the last well in which serum inhibited the hemagglutination of RBCs.

### QUANTIFICATION AND STATISTICAL ANALYSIS

All immune assay values were log10-transformed to improve linearity. Statistical significance was established at p<0.05. All reported p values are based on two-tailed tests. Correlation (Pearson), linear regression, local regression fit-line and related-sample comparison (Friedman’s two-way analysis of variance by ranks and pairwise comparison adjusted by Bonferroni correction) were performed using IBM SPSS Statistics (version 26).

## Notes

### Author Declarations

The study protocol was approved by the Institutional Review Board of University Hospital of Bellvitge, Barcelona, Spain; and by the Icahn School of Medicine at Mount Sinai, New York, US.

