## SupplementalFigures for "Antibody Immunological Imprinting on COVID-19 Patients"

A.

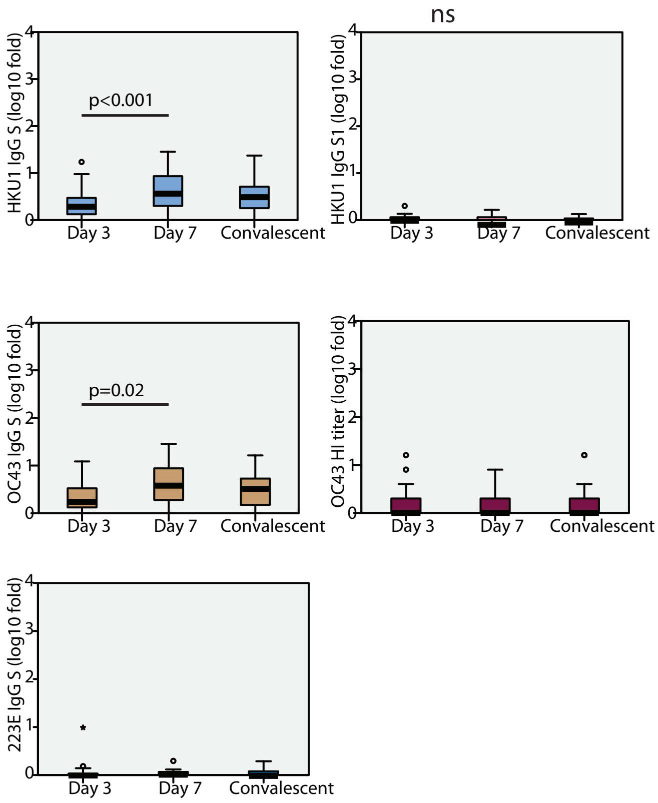


Supplemental Figure 1

Fig. S1. Fold induction of anti- human seasonal coronaviruses antibodies over time. A) Boxplot diagram of fold induction values of ELISA AUC titers against HKU1, OC43 and 223E at each time point: HKU1 IgG spike, HKU1 IgG S1, OC43 IgG spike and 223E IgG spike; and OC43 hemagglutination titers. Related-samples Friedman's two-way comparison was performed and significant adjusted p values for pairwise comparisons are shown for each antibody levels at each time point. Black bar indicated median values, box indicates IQR (Q1-Q3), lines indicate minimum and maximum. Outliers from the observed distribution are shown when present in each case.

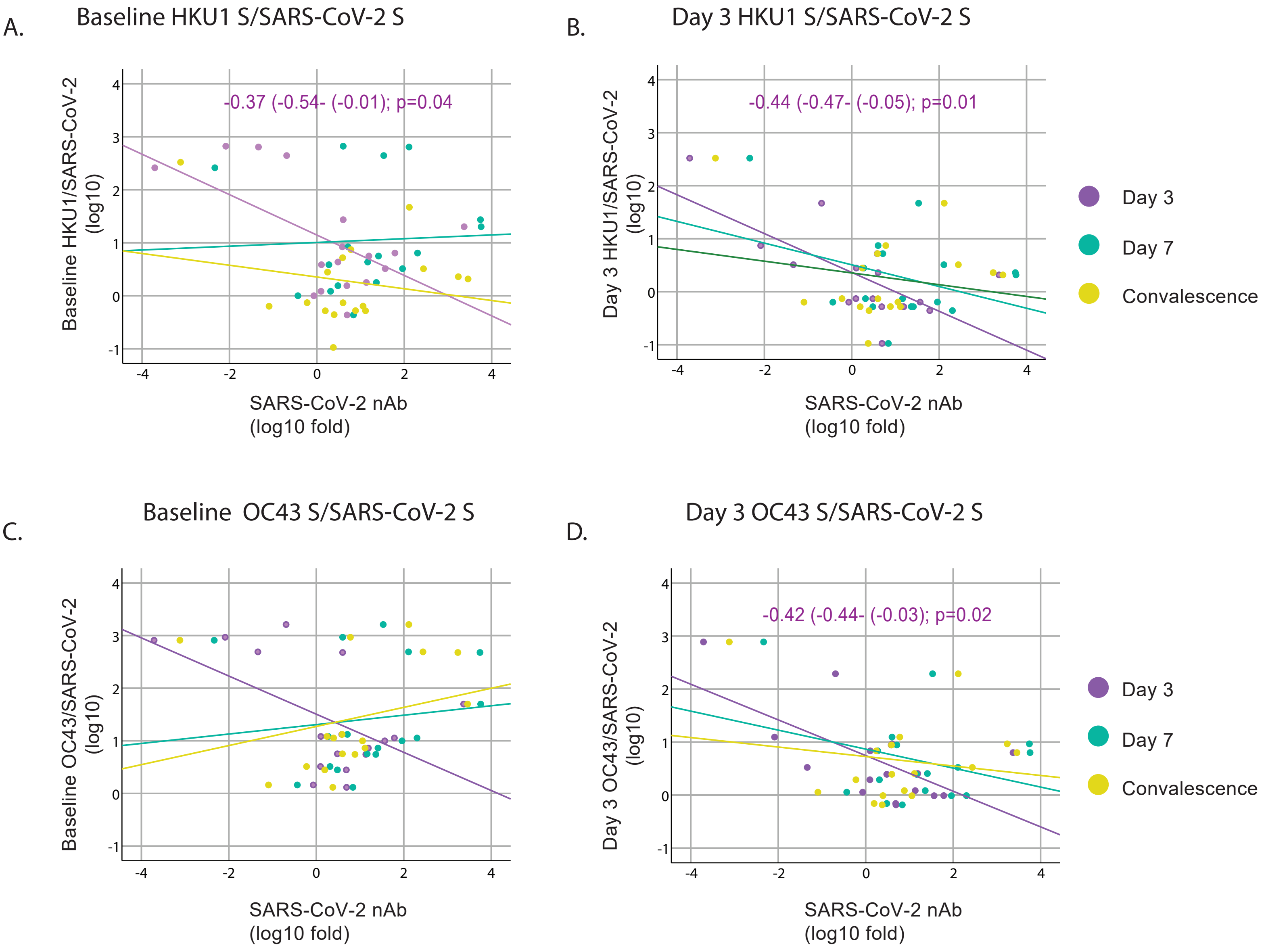


Fig S2. Influence of levels of back-boosting to HKU1 and OC43 normalized by levels of anti- SARS-CoV-2 on the induction of the novo neutralizing antibodies. A-D) Scatterplot of baseline and day 3 IgG levels for HKU1 and OC43 S protein normalized by the levels of SARS-CoV-2 IgG and their relationships with fold induction of SARS-CoV-2 neutralizing antibodies (nAb) over time. Overlay shows linear regression at each time point. Fitted linear regression and standardized beta coefficient (95% Confidence Interval, CI) for significant regression are shown.
